# The Prevalence and Management of Atrial Fibrillation in New Zealand Māori Detected through an Abdominal Aortic Aneurysm Screening Programme

**DOI:** 10.1101/2023.03.24.23287723

**Authors:** P. Sandiford, K. Poppe, C Grey, R. Doughty, E. Chambers, K.J. Kim, A. Hill, K. Bartholomew

## Abstract

**Background:** Atrial fibrillation (AF) screening was incorporated into an abdominal aortic aneurysm screening (AAA) programme for New Zealand (NZ) Māori.

**Methods:** AF screening was performed as an adjunct to AAA screening of Māori men aged 60-74 and women aged 65-74 registered with primary health care practices in Auckland, NZ. Pre-existing AF was determined through coded diagnoses or medications in the participant’s primary care record. Subsequent audit of the record assessed accuracy of pre-screening coding, medication use and clinical follow-up.

**Results:** Among 1955 people screened, the prevalence of AF was 144 (7.4%), of which 46 (2.4% of the cohort) were patients without AF coded in the medical record. More than half of these were revealed to be known AF but that was not coded. Thus, the true prevalence of newly detected AF was 1.1% (n=21). An additional 48 (2.5%) of the cohort had been coded as AF but were not in AF at the time of screening. Among the 19 at-risk screen-detected people with AF, 10 started appropriate anticoagulation within 6 months. Of the 9 who did not commence anticoagulation, five had a subsequent adverse clinical outcome in the follow-up period, including one with ischaemic stroke; two had contraindications to anticoagulants. Among those with previously diagnosed AF, the proportion receiving anticoagulation rose from 57% pre-screening to 83% at 6 months post-screening (p<0.0001); among new AF the proportion rose from 0% to 53% (p<0.01).

**Conclusions:** There is a high prevalence of AF in NZ Māori. AF screening is a feasible low-cost adjunct to AAA screening with potential to detect previously undiagnosed AF and reduce ethnic inequities in stroke. It may also prompt better coding and management of previously diagnosed AF. However, effective fail-safe measures are needed to ensure that high-risk newly diagnosed AF is managed according to best practice guidelines.

## Introduction

In Western Europe, North America and Australia 17% of ischaemic stroke is attributable to atrial fibrillation (AF).^1^ Strokes associated with AF tend to be severe with high disability and are prone to recurrence. Anticoagulants are highly effective in both primary and secondary stroke prevention, with novel oral anticoagulants (NOACs) such as apixaban and dabigatran superior and safer to warfarin.^2,3^ Their use is now considered best practice for the management of AF.^4,5^ and may be contributing to declining mortality rates from stroke in high-income countries,^6^ However the health impact depends upon accurate and timely detection of AF in those at high risk.

One limitation of NOAC stroke prophylaxis is that AF is often asymptomatic and/or paroxysmal leaving a significant proportion of cases undiagnosed and therefore unprotected. Disparities in health service access also affects diagnosis of both symptomatic and asymptomatic AF leading to inequities in stroke incidence and anticoagulant prophylaxis.^7^ Although the efficacy of AF screening for the prevention of stroke has not been demonstrated in randomised controlled trials,^8^ a recent review concluded that “*Screen-detected AF as found on single-timepoint screening or intermittent 30-second recordings over 2 weeks is not a benign condition and, with additional stroke risk factors, carries sufficient risk of stroke to justify consideration of screening and therapy to prevent stroke*.*”*^8^

The indigenous Māori population of Aotearoa New Zealand (NZ) is disproportionately impacted by ischaemic stroke compared to the European ethnicity population, with higher incidence and mortality rates, and lower survival.^7,9-11^ However little is known about the risk factors driving these health inequities.

In common with other indigenous groups, NZ Māori have a significantly higher age-specific prevalence of diagnosed AF than non-Māori,^12-14^ but studies have not determined the prevalence of undiagnosed AF in either group. Māori with diagnosed AF are not less likely to receive anticoagulation,^5^ highlighting the potential importance of undiagnosed AF in driving inequities in stroke incidence. Identifying factors relevant to AF screening for indigenous populations is a public health priority.^15^

Several screening strategies for asymptomatic undiagnosed AF have been proposed, and many different diagnostic modalities are now available, including inexpensive single lead mobile diagnostic tools which have made this more feasible.^8^ Assuming treatment of screen-detected AF is efficacious, opportunistic screening at health facilities is probably more cost-effective than a stand-alone screening programme,^16^ but inequities may be exacerbated more by opportunistic than organised screening.^17^ Organised AF screening could be of similar or higher cost-effectiveness to opportunistic screening if incorporated within an existing population-based programme such as influenza immunisation or abdominal aortic aneurysm (AAA) screening, rather than as a stand-alone vertical programme.^18^ Here we explore the potential of AF screening as an adjunct to an AAA screening programme for Māori in Auckland, NZ with the aim of improving equity as well as health outcomes.

## Methods

Screening for AF was incorporated as an adjunct within a pilot programme for AAA screening in Auckland NZ, the details of which have been described previously.^19^ The programme was the first targeted screening programme developed and tailored specifically to address inequities in AAA mortality.^20^ Adaptations include strong Māori leadership, partnership with Iwi (tribal) organisations, initiation with community champions, Māori programme staff and centralisation of whānau (family) experience. The study population included Māori men aged 60-74 and Māori women aged 65-74 registered with a primary care practice within the boundaries of Waitematā and Auckland District Health Boards. Although lower than non-Māori, over 90% of Māori aged 65 and over are registered with a primary care provider in NZ.^21^ Invitation to screening was made by a posted letter and information sheet (with translated versions available) followed up with a phone call.

Ethnicity data protocols in NZ require self-identification and allow multiple ethnicities to be recorded.^22^ A ‘prioritised’ ethnicity protocol was employed whereby a person having any of their ethnicities recorded as was classified as Māori and therefore eligible to participate. The quality of health sector ethnicity data in NZ is, if not perfect, generally very good.

AF screening was performed using an AliveCor^®^ KardiaMobile device which produces a single lead ECG interpreted by a paired smartphone app as either normal, AF, bradycardia or tachycardia, or unclassified. Participants found to be in AF were told that their results would be sent to their GP and to follow up with their doctor. Recordings not interpreted as ‘normal’ were sent to a cardiac physiologist (KP) for interpretation and reporting back to the GP using electronic messaging integrated with the practice management system. The message to the GP incorporated links to AF best practice resources and details of recommended anticoagulants, with a request to consider treatment after seeing the patient and performing a 12-lead ECG. GPs could also refer the patient for cardiology assessment. Where AF at the time of screening could not be ruled out due to recording quality or heart rate, this was stated in the GP message and it also advised a 12-lead ECG should be conducted to confirm the rhythm if clinically indicated. Co-payment of the primary care follow-up consultation was funded by the screening programme. A sample of normal ECG recordings were also checked by the cardiac physiologist.

In addition to AF and AAA screening, the visit included blood pressure measurement and for current smokers, a brief quit advice intervention with an offer of referral to a smoking cessation service. After the screening project was completed, an audit of patient’s primary care records was performed for all patients with AF (previously documented and newly diagnosed) or who remained unclassified after screening, to determine coding accuracy, CHA_2_DS-VASc score (an indicator of the 1-year risk of a thromboembolic event in patients with AF), prescribed treatments and any subsequent AF related events.

## Results

Of the 1979 people who attended AAA screening, 24 were not screened for AF as they had a pacemaker in situ leaving 1955 people in the cohort for AF screening. Of these, the expert reader was unable to rule out AF in 22 recordings reported as Unclassified by the AliveCor device, 1789 were confirmed as not having AF, and 144 people had confirmed AF at screening (**Table** 1). The prevalence of screen-detected AF was significantly higher in males than females (9.0 vs 5.0%; p<0.001).

**Table 1.**
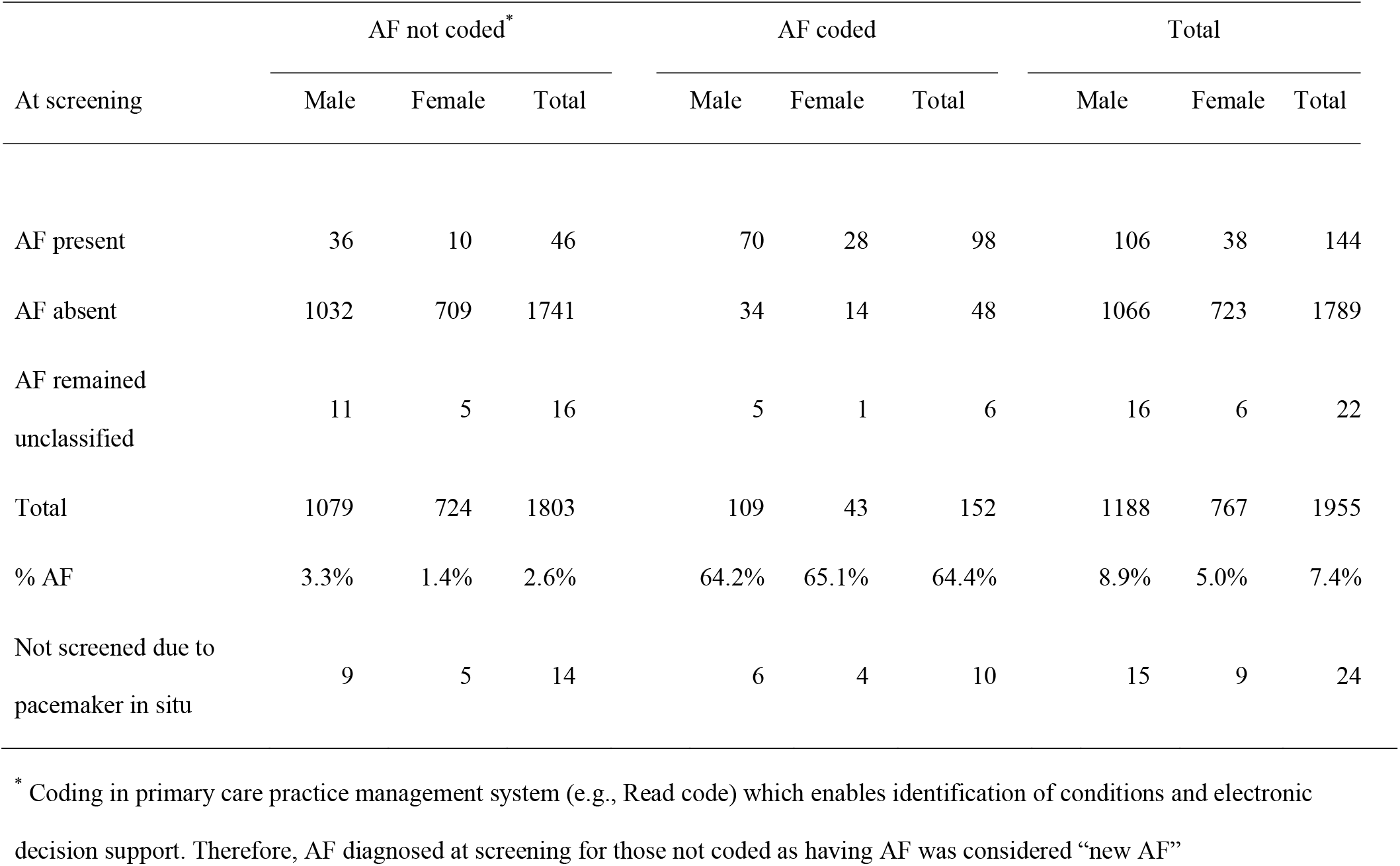
Results of AF screening in Māori attending AAA screening

Over one third of those with AF confirmed at screening (n=46, 2.4% of cohort) did not have a coded AF diagnosis in the electronic medical record. Subsequent audit revealed that over half of these (25/46) had been previously diagnosed, e.g. at an Emergency Department visit, but were either not coded or miscoded in the primary care medical record (**Table 2**). Among the 21 new screen-detected people with AF were 3 who were not in AF on a 12-lead ECG recorded later in primary care, and therefore were presumed to be paroxysmal AF.

**Table 2.** Results from subsequent audit of primary care medical records in those with previously coded or newly detected AF, or who remained unclassified after screening.

| In the primary care medical record |  |  |  |
| --- | --- | --- | --- |
|  | AF Coded<br>and Diagnosed | AF Not Coded<br>but Diagnosed | AF Not Coded<br>and Not Diagnosed |
| **In AF at screening** |  |  |  |
| N | 98 | 25 | 21 |
| CHA<sub>2</sub>DS-VASc≥2 | 88 (90%) | 23 (92%) | 19 (90%) |
| Anticoagulated |  |  |  |
| ▪ Pre-screening | 60 (68%) | 9 (39%) | 0 (0%) |
| ▪ Post-screening | 82 (93%) | 20 (87%) | 10 (53%) |
| **Not in AF at screening** |  |  |  |
| N | 46 | Did not meet criteria for audit |  |
| CHA<sub>2</sub>DS-VASc≥2 | 42 (91%) | | |
| Anticoagulated |  |  |  |
| ▪ Pre-screening | 17 (43%) |  |  |
| ▪ Post-screening | 24 (57%) |  |  |
| **Unclassified at screening** |  |  |  |
| N | 6 | 0 | 14 |
| CHA<sub>2</sub>DS-VASc≥2 | 5 (83%) | - | 11 (79%) |
| Anticoagulated |  |  |  |
| ▪ Pre-screening | 3 (60%) | - | 1 (7%) |
| ▪ Post-screening | 5 (100%) | - | 0 (0%) |

Six of the 22 with uninterpretable recordings had previously diagnosed and coded AF. The remaining 16 people were categorised as unknown AF status and have been excluded from calculation of AF prevalence. The prevalence of AF in the cohort is therefore the total of new screen-detected AF (n=21) plus known coded AF (n=152) plus known uncoded AF (n=30), out of people able to be screened for AF excluding those with unknown AF status (n=1955), equating to 10.4% — **Figure 1**).

**Figure 1.**
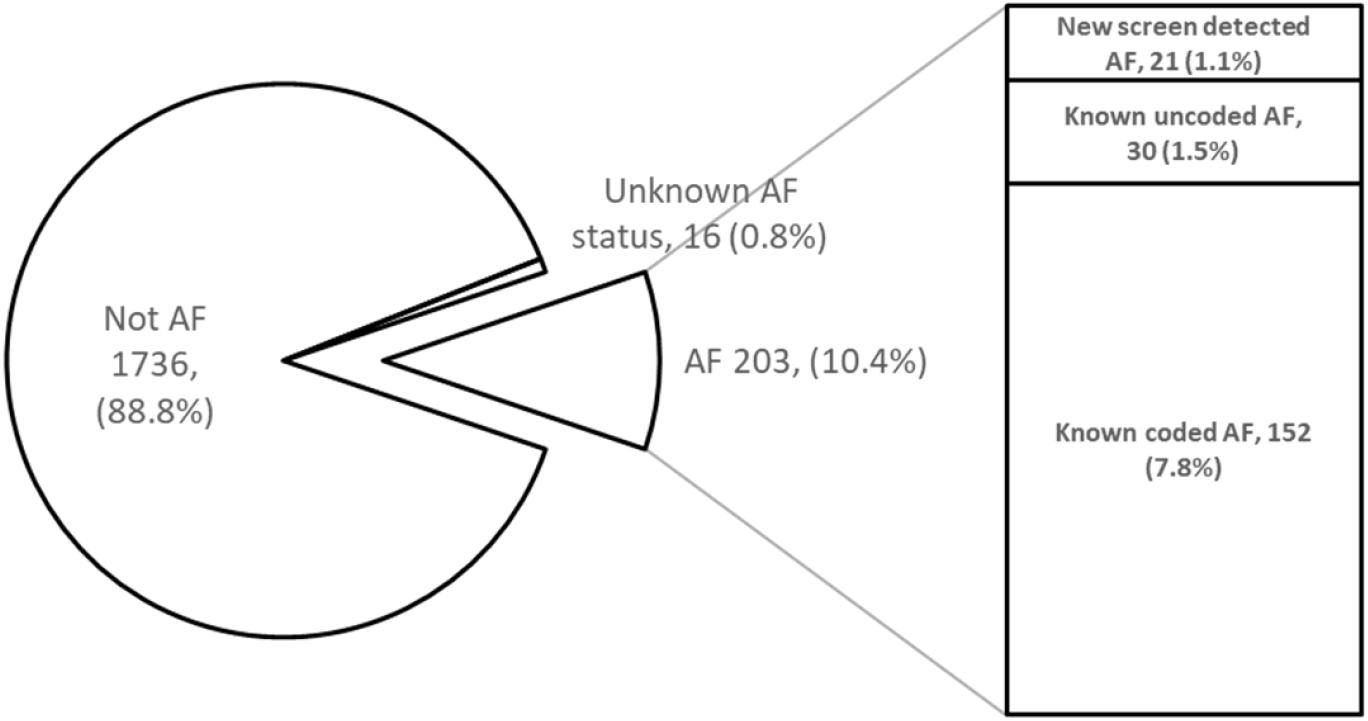
Prevalence of AF in screened population, by category

Records of 4 patients were unavailable for audit: two were AF coded but not in AF at screening; two did not have AF coded and the screening ECG was unable to be classified. At audit, the CHA_2_DS_2_-VASc scores were high for all people who had AF either detected at screening or previously known, with around 90% having a score of 2 or more. Only 57% (91/161) of patients with known AF (coded or not) and CHA_2_DS_2_-VASc scores of 2 or more (referred to henceforth as ‘at risk’) were on anticoagulation prophylaxis prior to screening, but a significantly higher proportion (83%, 134/161; p<0.0001) were anticoagulated at 6 months following screening, suggesting that screening may have led to better prophylactic management. The change was also significant in the 19 at risk newly diagnosed AF group (0% pre-screening vs 53% post-screening; p<0.01). Of 9 at-risk patients with newly diagnosed AF that were not started on appropriate anticoagulation, 5 (56%) had a subsequent adverse clinical outcome. All 5 either did not get followed-up by their GP or were seen but no AF discussion was recorded: one with a CHA_2_DS_2_-VASc score of 4 subsequently had an ischaemic stroke 6 weeks post-screening; two had subsequent inpatient hospital admissions with “new fast AF” 2 and 4 years post-screening; one was admitted a year after screening with venous thrombosis; and one had an inpatient hospital admission 3 months post-screening with venous thrombosis and “new fast AF”. Two of the 9 cases not on anticoagulation prophylaxis had recorded contraindications and were taking aspirin, three were no longer in AF when the practice repeated the ECG.

Receiving anticoagulants was more common in those at risk with coded AF or known uncoded AF who were in AF at the time of screening, than in those who were not in AF at screening, both before screening (62% vs 42%) and after screening (92% vs 60%). Apart from the lack of follow-up, other reasons identified at audit for not prescribing anticoagulants included patient contraindications, cardiologist recommendation, successful ablation treatment and patient choice.

## Discussion

Our study has demonstrated the feasibility of including AF screening as a co-benefit of AAA screening, which should increase the cost-effectiveness of both. AF could be an adjunct to any cardio-vascular screening episode. To our knowledge this is the first community-based assessment of the prevalence of diagnosed and undiagnosed AF among Māori in NZ. Previous research has used electronic medical records to quantify prevalence based on recorded diagnoses but AF was not systematically tested for in that study population.^23^ In our population the proportion with a pre-existing diagnosis of AF was 7.8% (152/1955) which was the same as that in Gu *et al*’s study of Māori in primary care aged 60-74 years (128/1620, 7.9%).^23^ If the miscoded/uncoded cases are included, the prevalence of AF rises to 9.1% (177/1955) in our study; adding truly new cases, the prevalence is 10.5%. The prevalence in Gu *et al’s* non-Māori non-Pacific population of the same age range was just 3.8%, underlining the ethnic inequity in AF.^23^

Despite the high overall prevalence of AF in this population, only 1.1% were newly diagnosed at screening. This may call into question the cost-effectiveness of AF screening in this population but the low marginal cost of the programme when implemented as an adjunct to AAA screening stands in its favour. Also, the marked increase in anticoagulation prophylaxis observed among known and uncoded AF patients is an important additional impact of the programme. A rigorous cost-effectiveness analysis would be helpful.

The effectiveness of AF screening in stroke prevention depends upon high-risk patients receiving oral anticoagulant prophylaxis when AF is detected. Perhaps the most striking finding of this study is the frequently inadequate response to a diagnosis of AF, the reasons for which are not entirely clear. Our findings suggest a combination of communication break-down between the GP, practice administrative staff and patients, in organising follow-up reviews, as well as lack of knowledge on the latest guidelines for AF management. But given that 95% of Māori aged 65-74 have CHA_2_DS_2_-VASc scores of 2 or more,^24^ it seems reasonable to expect AF to be treated more aggressively than was observed, and this was underlined by the high frequency of subsequent adverse outcomes in the at-risk newly diagnosed patients who did not receive anticoagulation. Our programme has changed practice to incorporate a failsafe follow up at 4 months post-diagnosis for all new AF, with prompts to primary care to recall the patient. Other improvements to primary care management are underway in New Zealand, including data matching with secondary care to improve AF coding, improved electronic decision support and better linkage with cardiovascular risk assessment.^25^

The contribution of AF to ethnic differences in health outcomes depends on two factors: prompt and accurate diagnosis of symptomatic and asymptomatic AF; and treating those at high-risk with appropriate anticoagulants. While our study led to anticoagulant prophylaxis in a relatively small number of at-risk Māori with previously undiagnosed AF (n=10), a much larger number of at-risk Māori with known AF (n=43) commenced anticoagulant treatment in the 6 months following screening which we assume was in response to the feedback provided to primary care practices by our programme. Although Māori are as likely or more likely than non-Māori to receive anticoagulation,^5,24^ the much higher prevalence of AF among Māori means that the impact of inadequate management of known AF will disproportionately affect health outcomes for Māori and contribute to ethnic inequities due to stroke. The inequity may be partly perpetuated by poor coding if these patients remain ‘hidden’ from clinical efforts to improve AF management.

It was noted that a high proportion of patients previously diagnosed with AF were in sinus rhythm at the time of screening. It is presumed that most of these were cases of paroxysmal AF and not misdiagnoses but that is difficult to demonstrate. Equally we may have missed diagnosing patients with paroxysmal AF if they were in sinus rhythm during the screening.

In conclusion, the study has shown that previously undiagnosed AF can be detected at AAA screening and that it may have a significant impact on improving guideline-indicated management of patients with known AF. Given the relatively low marginal cost, these co-benefits and the impact on inequity may justify the resources used. What the study has also revealed is that even with a robust system for informing patients and primary care when new cases of AF are detected at screening, many still receive inadequate management requiring safety net follow up by the programme. Optimising AF management once AF is identified needs to be addressed before the benefits of AF screening will lead to systematic reductions in health inequities.

## Data Availability

Data is available on request to the corresponding author

## Acknowledgments

The authors acknowledge the hard work of Anna Maxwell and Mellissa Murray in the AAA pilot which has provided valuable inputs for the preparation of this manuscript. Also, Matua (elders) Rahiri Bennett and Fraser Toi as the project’s cultural champions. Ethical approval for the project was obtained from the Northern B Health and Disability Ethics Committee on 12/3/2015 (ref 15/NTB/AM03) which was extended in 2017 to include Auckland district participants. The screening programme funding was provided by the Auckland and Waitemata District Health Boards. KKP was supported by a Heart Foundation of New Zealand Hynds Senior Fellowship at the time of this study.

